# Distinct circulating proteomic patterns associated with thoracic aortic diameter and stiffness: A proteome-wide study in the UK Biobank

**DOI:** 10.64898/2026.11.13.26368064

**Authors:** Andy Choi, Adrian J. Hobbs, Janice Tsui, Aroon D. Hingorani, Nikhil Paliwal

## Abstract

**Background:** Thoracic aortic aneurysm (TAA) is characterized by abnormal dilation and functional remodelling of the thoracic aorta. Current detection and surveillance rely largely on imaging, limiting population-wide risk stratification. Circulating proteins may provide biological insight into aortic remodelling and support future biomarker development. We aimed to identify circulating proteomic patterns associated with imaging-derived thoracic aortic diameter and aortic stiffness in a large population-based cohort.

**Methods:** A proteome-wide association study was conducted in 6773 UK Biobank participants with available plasma proteomic and cardiac imaging data. A combined thoracic aortic diameter was derived from taking the maximum of standardized ascending and descending measurements, while stiffness was derived using a pressure-free index of aortic wall properties. Associations with 2920 circulating proteins were evaluated using multivariable linear regression with Benjamini-Hochberg false discovery rate (FDR) correction. Functional enrichment analysis was performed using the complete ranked protein list across Hallmark, curated, and Gene Ontology Biological Process collections. Principal component analysis (PCA) was performed as a secondary analysis to explore shared and divergent variation between the two phenotypes.

**Results:** In age- and sex-adjusted analyses, 358 proteins were associated with aortic diameter and 60 with stiffness after FDR correction. Sequential adjustment materially altered these associations; in the primary fully adjusted models, 417 proteins were associated with diameter and six with stiffness. Functional enrichment analyses highlighted complement/coagulation, extracellular matrix organization, and contractile pathways associated with aortic diameter. In contrast, although only a few individual proteins were associated with stiffness, rank-based pathway analysis across the full proteome identified a distinct enrichment pattern involving cytokine signalling, protein processing, and intracellular transport. Aortic diameter and stiffness were only modestly correlated (r = 0.21). Secondary PCA showed that the first principal component (PC1) largely overlapped with the diameter-associated profile, with 310 of 370 PC1-associated proteins also associated with diameter. Four proteins (*REN, BCAN, GGH and THY1*) were associated with both diameter and stiffness in addition to being associated with PC1.

**Conclusion:** Circulating proteomic profiling identified markedly different and minimally overlapping molecular patterns associated with thoracic aortic diameter and stiffness. Structural dilation was primarily linked to complement/coagulation, extracellular matrix, and contractile pathways, whereas stiffness reflected a smaller, more distinct aspect of aortic remodelling involving cytokine signalling and cellular transport processes. The four shared proteins across both phenotypes (*REN, BCAN, GGH* and *THY1)* represent candidates for further biomarker and causal investigation.

## Introduction

Thoracic aortic aneurysm (TAA) is a life-threatening vascular condition characterized by the abnormal focal dilation of the thoracic aorta, encompassing the aortic root, ascending aorta, aortic arch, and descending aorta. TAAs rarely manifest with symptoms but can continue to enlarge until they rupture, at which point the mortality can exceed 90-97% without immediate intervention due to fatal haemorrhage (1). It is estimated that TAA affects around 5.3 per 100,000 individuals and around 1.6 per 100,000 individuals in the general population experience a rupture each year (2).

In several healthcare systems, including the National Health Service (NHS) in the UK, abdominal aortic aneurysm (AAA) has an effective screening programme currently available to men ≥ 65 years, which has significantly reduced mortality and associated costs; however, TAA lacks any similar option (3–5). This partly reflects the need for more resource-intensive imaging modalities, such as computed tomography (CT) or magnetic resonance imaging (MRI) (6,7). Due to the lower prevalence of TAA in the general population, current surveillance is largely reserved for individuals with a strong family history, a known genetic syndrome (i.e. Marfan syndrome or Ehlers-Danlos syndrome), specific pathological variants (i.e. *ACTA2*, *MYH11*, *COL3A1*), or incidentally detected thoracic aortic dilation (4,5). In non-genetic cases, thoracic aortic diameter has been proposed for surveillance and intervention (4,5).

Beyond clinically diagnosed aneurysms, quantitative measures of diameter and stiffness of the ascending and descending thoracic aorta may provide insight into earlier stages of the disease. Aortic diameter reflects the structural dilation and remains the principal measure used for clinical surveillance and intervention decisions (4,5). In contrast, aortic stiffness captures functional changes that may occur prior to or during vascular remodelling, associated with vascular ageing and consequently adverse cardiovascular outcomes (8). Importantly, aortic stiffness may occur prior to aortic dilation and has been investigated for its role in aortic dissection among patients with a dilated ascending aorta (9,10). Examining these traits together therefore can help identify shared and phenotype-specific proteomic associations with thoracic aortic remodelling.

Large-scale plasma proteomics profiling provides an opportunity to investigate circulating biomarkers and explore enriched pathways associated with the underlying vascular pathology in large population-based cohorts (11,12). Previous studies leveraging the UK Biobank data have identified genetic determinants of TAA that may improve prediction (13,14). Additionally, recent studies have separately examined plasma proteomic correlates associated with aortic diameter or stiffness in both large and small patient cohorts; however, the specific biology of how dilation and stiffness are related to TAA remains poorly understood (15,16). It also remains unclear whether the correlates of thoracic aortic dilation overlap with, or differ from, those from thoracic aortic stiffness.

Therefore, this study was designed to analyse the extensive proteomics data and imaging-derived aortic phenotypes held in the UK Biobank to identify biologically relevant associations between circulating proteins and thoracic aortic diameter and/or stiffness. By also taking into consideration the overlap between proteins associated with structural dilation and functional stiffening, we aimed to distinguish between shared and phenotype-specific proteomic signatures of thoracic aortic remodelling.

## Methods

### Study population

The UK Biobank is a population-based cohort of over 500,000 volunteers, aged 40-69 at baseline (predominantly of self-reported white ethnicity), who were recruited between 2006 and 2010 (17). For the present study, the initial pool consisted of 53,058 participants randomly invited for plasma proteomic profiling at the baseline visit and 56,534 individuals who underwent cardiac magnetic resonance imaging (CMR). Only those with both plasma proteomics and CMR data, a total of 6,773 participants, were analysed in the main cohort, as detailed in Fig. 1. All participants had complete covariate data available.

**Figure 1:**
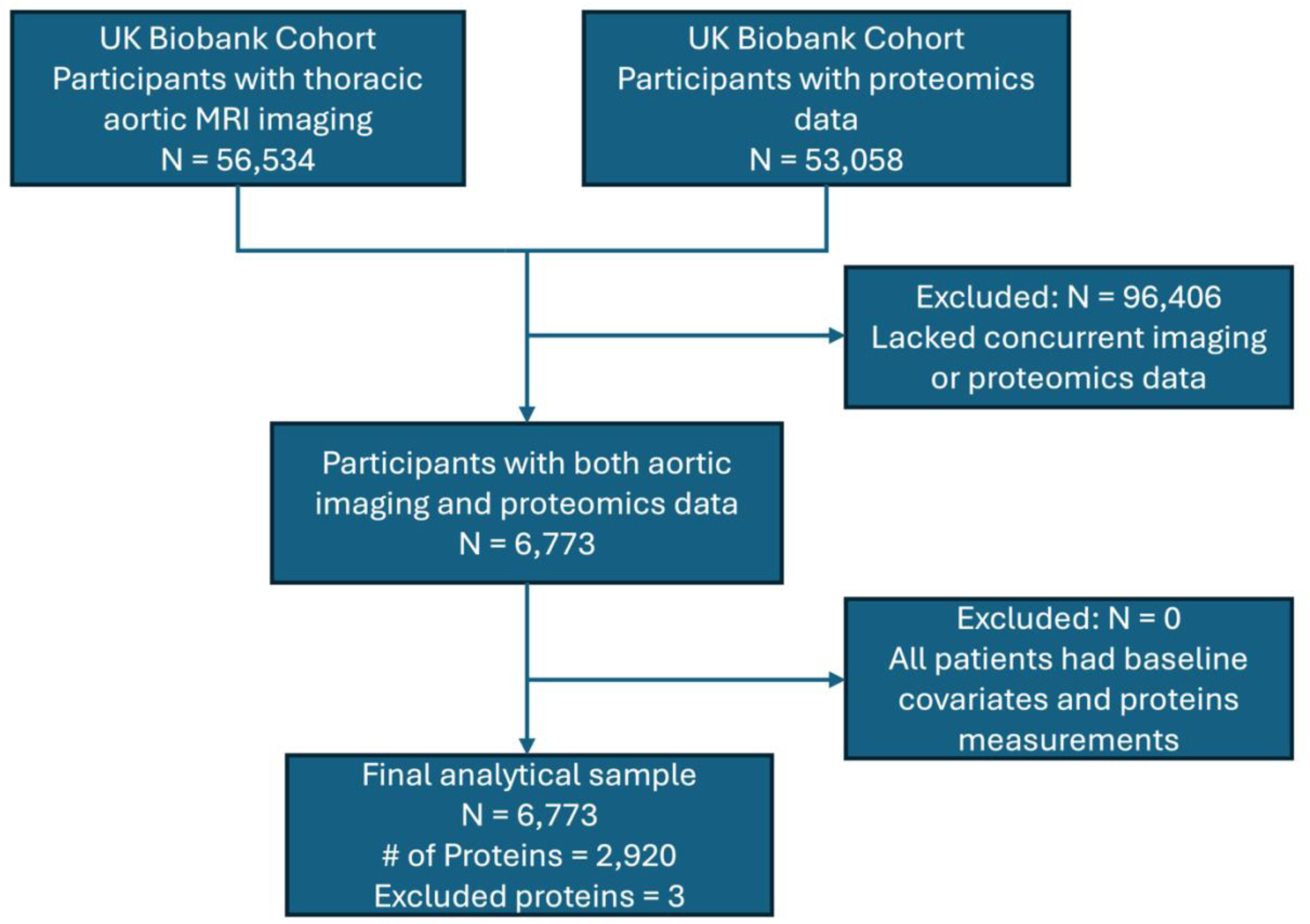
Participant flow pipeline for the UK Biobank participants included, depicting the recruitment, exclusion criteria, and final sample selection process. Final analytical cohort (N = 6773) was evaluated against 2920 circulating proteins after quality control filtering and selecting for participants with overlapping cardiovascular magnetic resonance imaging and Olink plasma proteomic data.

The UK Biobank CMR procedure has previously been detailed (18–20). During an imaging visit, CMR imaging is taken alongside a carotid ultrasound, a brain MRI scan, and a dual energy X-ray absorptiometry (DEXA) scan (20). Each CMR image was then automatically segmented by applying an in-house pre-trained neural network to identify the aorta. Bai et al. further describe the methodology behind the neural network model (19). The maximum aortic area was subsequently identified and used for analysis with the assumption of a circular aortic cross-section (21).

Participant data included genetic, imaging, physical and anthropometric measurements, linked health electronic health records, and proteins in the blood. The proteomics profiling was performed using the antibody-based Olink Explore Proximity Extension Assay (PEA), which included eight protein panels: cardiometabolic, cardiometabolic II, inflammation, inflammation II, neurology, neurology II, oncology, and oncology II. The UK Biobank was approved by the North West Research Ethics Committee (21/NW/0157), and all participants provided informed consent at recruitment.

Baseline characteristics were collected during the first assessment visit. The covariates used in the analysis include age, sex, body mass index (BMI), systolic blood pressure (SBP), smoking status, and ethnicity. Plasma proteomic profiling and cardiac imaging were performed at different visits from the initial visit, reflecting the UK Biobank’s staged recruitment design (17). As with other UK Biobank proteomics studies, a temporal separation was seen between proteomic sampling and imaging acquisition (22–24).

### Aortic imaging phenotypes

To account for anatomical variations, raw ascending and descending thoracic aortic diameters were separately scaled and transformed into standard Z-scores, calculated as Z = (x-µ)/*σ*, where (x) represents the individual participant value and (µ) and (*σ*) represent the reference mean and standard deviation of participants with available aortic imaging phenotypes, prior to restricting to only those with available proteomics data (21). Hence, a unit increase in the Z-score corresponds to a 1-standard-deviation increase in the aortic phenotype. A combined thoracic aortic diameter phenotype was defined as the maximum of the ascending and descending diameter Z-scores to capture the peak regional abnormality from the imaging reference population.

Aortic stiffness was quantified using a pressure-independent index, which was calculated from ascending aortic diameter obtained from aortic CMR imaging together with the systolic and diastolic blood pressure measurements (25). The measure assumes an exponential relationship between pressure and diameter, and the resulting measure was similarly normalized via Z-score transformation (21,25). Distributions of the diameter and stiffness phenotypes were assessed using histograms and quantile-quantile plots to verify that they followed an approximately Gaussian distribution prior to regression analysis.

### Primary proteome-wide association analyses

The primary analysis of the thoracic aortic phenotype was evaluated across two levels: thoracic aortic diameter and aortic stiffness. Their association with circulating protein levels was assessed using multivariable linear regression models implemented through the “data.table” R package (26). Protein measurements were standardized within each protein target group to account for cross-protein effect comparisons.

To assess proteomic associations with the respective aortic phenotype, sequential covariate-adjustment models were fitted to assess the extent to which BMI and SBP influenced protein-aortic phenotype associations. The adjustments for age and sex were considered the baseline adjustment set given the well-established associations with both circulating protein levels (27,28). For each phenotype, Model 1 was adjusted for age and sex; Model 2 was adjusted for age, sex and BMI; Model 3 was adjusted for age, sex, and SBP; Model 4 included age, sex, BMI and SBP; and Model 5 included further adjustments for smoking status and ethnicity. This sequential approach was used to evaluate changes in the magnitude of protein-phenotype associations following adjustment for BMI and SBP individually and in combination.

For aortic stiffness, SBP-adjusted models were interpreted as sensitivity analyses given that blood pressure is directly involved in the calculation of the stiffness phenotype. Therefore, an additional fully adjusted stiffness model (Model 6) was fitted to include age, sex, BMI, smoking status and ethnicity to prevent potential overadjustment. The false discovery rate (FDR) was accounted for by adjusting raw p-values across all models according to the Benjamini-Hochberg (BH) method (29). Results were considered statistically significant at an FDR-adjusted p-value < 0.05.

### Secondary PCA-derived analysis

As an exploratory analysis, a principal component analysis (PCA) was performed on a unified covariance matrix including both normalized stiffness and combined diameter measurements using singular value decomposition. The correlation between aortic diameter and stiffness was approximately 0.21, indicating a modest correlation between the two phenotypes. 1^st^ principal component (PC1) was interpreted as a shared-direction component of diameter and stiffness while 2^nd^ principal component (PC2) represents their divergent component. To ensure interpretability, the loading vectors were signed relative to the increasing values of aortic diameter.

Proteome-wide associations using sequential covariate-adjusted models were repeated for PC1 and PC2 as secondary analyses using the same multivariable framework as aortic diameter associations, adjusting for age, sex, ethnicity, BMI, SBP, and smoking status in the fully adjusted model. PC2 results were considered exploratory.

### Functional enrichment analysis

To gain biological insight into the functions of the proteins driving TAA phenotypes, Gene Set Enrichment Analysis (GSEA) and Over-Representation Analysis (ORA) were performed using the “clusterProfiler” R package (30). For GSEA, proteins were ranked continuously by a composite score, which was defined as: Rank = β / SE, where β is the multivariable regression coefficient from the primary fully adjusted model (Model 5 for aortic diameter and Model 6 for stiffness) and SE is the standard error. The ranked list was subsequently enriched according to the MSigDB Hallmark (H), Curated Gene Sets (C2), and Biological Processes (C5) human gene set collections using 10,000 permutations. Multiple-testing correction was applied separately within each phenotype and gene-set collection using the Benjamini-Hochberg method. This analysis was performed separately for each imaging phenotype.

The ORA was applied to evaluate discrete, threshold-based protein lists derived from phenotypic interactions. This included proteins significantly associated across both the diameter and PC1 phenotypes and proteins uniquely associated with individual phenotypes. These proteins were enriched according to Gene Ontology: Biological Processes (GO:BP) via the “org.Hs.eg.db” human annotations database (31). The ORA used successfully mapped Olink-tested proteins as the background universe. Enriched pathways with an FDR-adjusted p-value < 0.05 were selected. Protein-protein interaction analysis was performed as an exploratory follow-up for protein subsets showing significant ORA enrichment after FDR adjustment. This approach was used to visualize the relationships among enriched proteins.

### Additional secondary and sensitivity analyses

Given the significant role of extracellular matrix degradation in thoracic aortic aneurysm and the well-documented link between elastin (*ELN*) and thoracic aortic disease (4,5,32), a sensitivity analysis was conducted centred around the *ELN* levels in the cohort. Associations between circulating *ELN* and each aortic phenotype were sequentially fitted across three ordinary least squares (OLS) models for both the individual metrics (stiffness and diameter) and the combined phenotype based on PC1: 1) unadjusted model based on the raw, unadjusted association between *ELN* and phenotype; 2) age-adjusted model; 3) fully adjusted model, controlled for the full baseline variables.

Additionally, to determine whether proteomic associations with thoracic aortic phenotypes vary by biological sex, the associations were tested via sex-interaction testing. For each protein, an ANOVA-based likelihood ratio test (LRT) was used to compare between the main-effects model and an interactions model including the term Sex × Phenotype. Interaction p-values were adjusted according to the BH method. Sex-stratified multivariable linear regressions were executed separately within female and male subgroups, adjusting for age, ethnicity, BMI, SBP and smoking status in the diameter phenotype models and excluding SBP for the stiffness phenotype models. Subgroup p-values were independently FDR-adjusted using the BH method.

## Results

### Study population characteristics

The final cohort, restricted to participants with both proteomic and imaging data available yielded a study sample of n = 6,773. Baseline demographic, cardiovascular, and imaging characteristics are summarised in Table 1. Participants had a mean age of 54.2 years (SD 7.7), and 51.4% were female. Most participants were of white ethnicity (96.07%); hence, other ethnicities were grouped together for analysis. 59.2% of the participants had never smoked, and the mean body mass index and systolic blood pressure were 26.5 kg/m^2^ (SD 4.1) and 135.8 mmHg (SD 18.37), respectively.

**Table 1:** Baseline characteristics of the UK Biobank study cohort. Demographic, anthropometric, cardiovascular, and imaging characteristics are summarised for participants with both plasma proteomic and cardiac magnetic resonance imaging data (n = 6773). Continuous variables are expressed as mean (standard deviation), and discrete variables are presented as counts and percentages (n (%)).

| **Characteristic** | **Overall cohort, n = 6773** |
|---|---|
| Age, years | 54.20 (7.73) |
| Female sex | 3481 (51.4%) |
| Ethnicity |  |
| White | 6507 (96.07%) |
| Mixed | 52 (0.77%) |
| Asian | 75 (1.11%) |
| Black | 64 (0.94%) |
| Chinese | 11 (0.16%) |
| Other ethnic group | 45 (0.66%) |
| Prefer not to answer | 18 (0.27%) |
| Do not know | 1 (0.015%) |
| BMI, kg/m<sup>2</sup> | 26.49 (4.14) |
| Systolic blood pressure, mmHg | 135.78 (18.37) |
| Smoking status |  |
| Never | 4008 (59.18%) |
| Previous smoker | 2289 (33.80%) |
| Current smoker | 464 (6.85%) |
| Prefer not to say | 12 (0.18%) |
| Ascending aortic area, mm<sup>2</sup> | 857.07 (193.44) |
| Descending aortic area, mm<sup>2</sup> | 477.85 (100.05) |
| Combined diameter Z-score | 0.00 (0.99) |
| Aortic stiffness Z-score | 0.00 (0.99) |

Continuous variables are reported as mean (SD).
Discrete variables are reported as n (%).

**Table 2:** Sequential covariate-adjusted associations of top circulating proteins with thoracic aortic diameter. Proteins are ranked in the list according to leading associations from the age- and sex-adjusted baseline model (Model 1). Standardised effect sizes (beta) for the top ten proteins are shown across sequential covariate-adjustment models. Model 1 was adjusted for age and sex; Model 2 additionally included BMI; Model 3 included SBP instead of BMI; Model 4 included age, sex, BMI and SBP; and Model 5 further included smoking status and ethnicity. The beta value reflects changes in the standard deviation (SD) of protein level per 1 SD increase in aortic diameter phenotype. Values are represented as beta (95% CI). Statistical significance was defined as Benjamini-Hochberg FDR-adjusted p-value < 0.05. All proteins shown met the significance threshold across all sequential models.

| **Protein** | **M1** | **M2** | **M3** | **M4** | **M5** |
|---|---|---|---|---|---|
| Neurocan core protein (*NCAN*) | -0.22 (-0.24, -0.19) | -0.15 (-0.17, -0.12) | -0.21 (-0.24, -0.18) | -0.15 (-0.18, -0.12) | -0.14 (-0.16, -0.11) |
| Seizure 6-like protein (*SEZ6L*) | -0.20 (-0.22, -0.17) | -0.13 (-0.16, -0.10) | -0.19 (-0.22, -0.16) | -0.13 (-0.16, -0.10) | -0.12 (-0.15, -0.10) |
| Myelin-oligodendrocyte glycoprotein (*MOG*) | -0.18 (-0.20, -0.15) | -0.12 (-0.15, -0.09) | -0.17 (-0.20, -0.14) | -0.12 (-0.15, -0.09) | -0.12 (-0.15, -0.10) |
| NTRK-like SLIT and protein 1 (*SLITRK1*); | -0.18 (-0.22, -0.15) | -0.11 (-0.14, -0.08) | -0.18 (-0.21, -0.15) | -0.11 (-0.14, -0.08) | -0.10 (-0.13, -0.07) |
| WAP, Kazal, immunoglobulin, Kunitz and NTR domain-containing protein 2 (*WFIKKN2*) | -0.16 (-0.19, -0.13) | -0.07 (-0.10, -0.04) | -0.15 (-0.18, -0.13) | -0.07 (-0.10, -0.05) | -0.06 (-0.09, -0.03) |
| Brevican core protein (*BCAN*) | -0.16 (-0.18, -0.13) | -0.10 (-0.12, -0.07) | -0.15 (-0.18, -0.12) | -0.10 (-0.13, -0.07) | -0.09 (-0.12, -0.06) |
| Secretogranin-1 (*CHGB*) | -0.15 (-0.18, -0.12) | -0.09 (-0.11, -0.06) | -0.15 (-0.18, -0.13) | -0.09 (-0.12, -0.06) | -0.09 (-0.12, -0.07) |
| Leucine-rich repeat and transmembrane domain-containing protein 2 (*LRTM2*) | -0.17 (-0.20, -0.14) | -0.11 (-0.14, -0.08) | -0.17 (-0.20, -0.13) | -0.11 (-0.14, -0.08) | -0.11 (-0.14, -0.07) |
| Opticin (*OPTC*) | -0.14 (-0.17, -0.11) | -0.09 (-0.12, -0.07) | -0.14 (-0.17, -0.11) | -0.09 (-0.12, -0.07) | -0.09 (-0.12, -0.06) |
| Neurosecretory protein VGF (*VGF*) | -0.16 (-0.20, -0.13) | -0.12 (-0.15, -0.08) | -0.16 (-0.19, -0.13) | -0.12 (-0.15, -0.08) | -0.12 (-0.15, -0.08) |

**Table 3:**
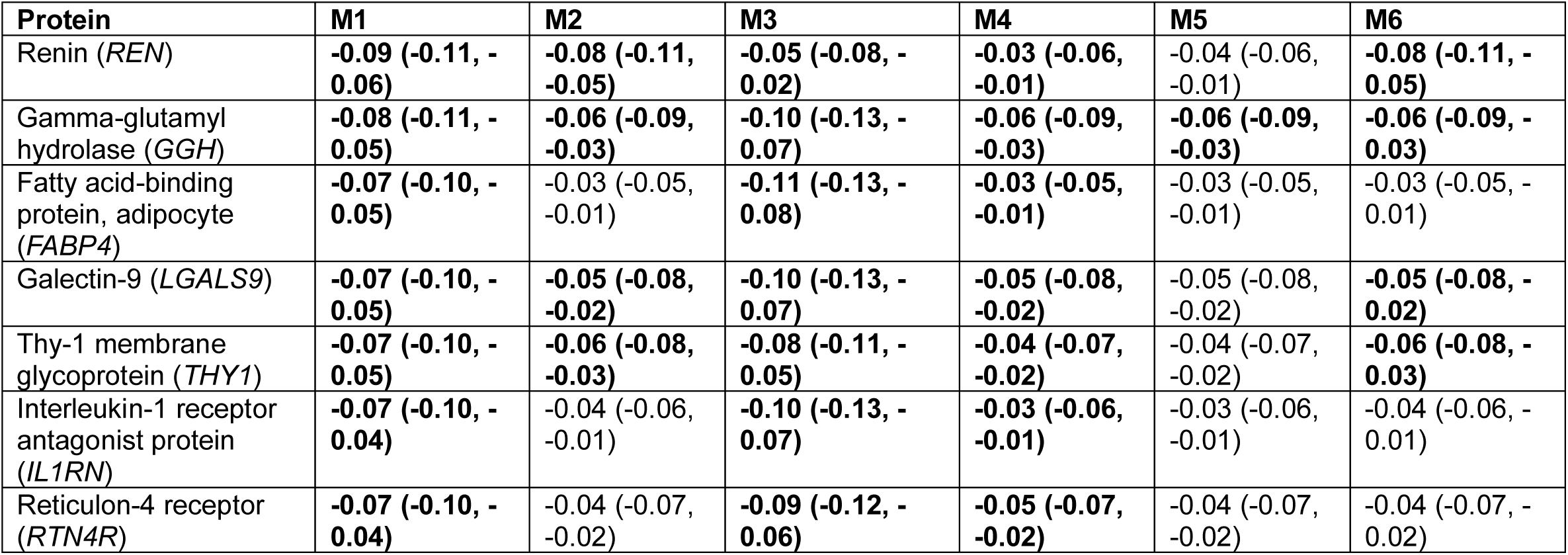
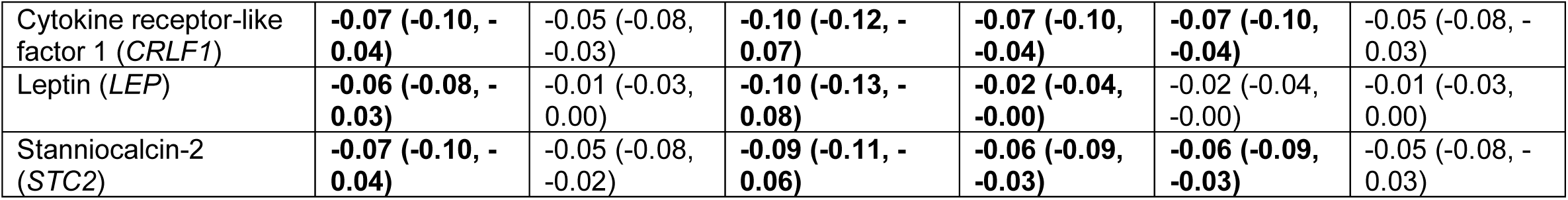
Sequential covariate-adjusted associations of top circulating proteins with aortic stiffness. Proteins are ranked in the list according to leading associations from the age- and sex-adjusted baseline model (Model 1). Standardised effect sizes (beta) for the top ten proteins are shown across sequential covariate-adjustment models. Model 1 was adjusted for age and sex; Model 2 additionally included BMI; Model 3 included SBP instead of BMI; Model 4 included age, sex, BMI and SBP; Model 5 further included smoking status and ethnicity; Model 6 adjusted for age, sex, BMI, smoking status and ethnicity without SBP. Because SBP contributes directly to the stiffness phenotypes, Models 3-5 were considered sensitivity analyses, while Model 6 represented the full primary analysis. The beta value reflects changes in the standard deviation (SD) of protein level per 1 SD increase in aortic stiffness phenotype. Values are represented as beta (95% CI). Statistical significance was defined as Benjamini-Hochberg FDR-adjusted p-value < 0.05. Bold values indicate associations meeting the FDR significance threshold.

Protein-level quality control was performed by assessing the coverage of the 2923 proteins across the entire cohort. Proteins from the core panels achieved near-complete cohort coverage (ranging from n = 5922 to n = 6670 observations), while other proteins were more restricted and had lower sample sizes in the cohort between n = 4339 and n = 5358. Three proteins (*GLIPR1*, *NPM1* and *PCOLCE*) were identified as having substantial missingness, with less than 50% of the cohort having available data, which has previously been documented in other UK Biobank studies (33). These proteins were therefore removed from downstream analyses, and a total of 2920 unique proteins were analysed.

### Proteome-wide associations with aortic diameter

In models adjusted for age and sex, 358 of 2920 circulating proteins were associated with thoracic aortic diameter following Benjamini-Hochberg FDR correction for multiple testing (Fig. 2a; Supplementary Table 1). The strongest associations included neurocan core protein (*NCAN)*; seizure 6-like protein (*SEZ6L)*; myelin-oligodendrocyte glycoprotein (*MOG)*; NTRK-like SLIT and protein 1 (*SLITRK1*); and WAP, Kazal, immunoglobulin, Kunitz and NTR domain-containing protein 2 (*WFIKKN2*). An inverse relationship was observed for the majority of proteins (n = 210).

**Figure 2:**
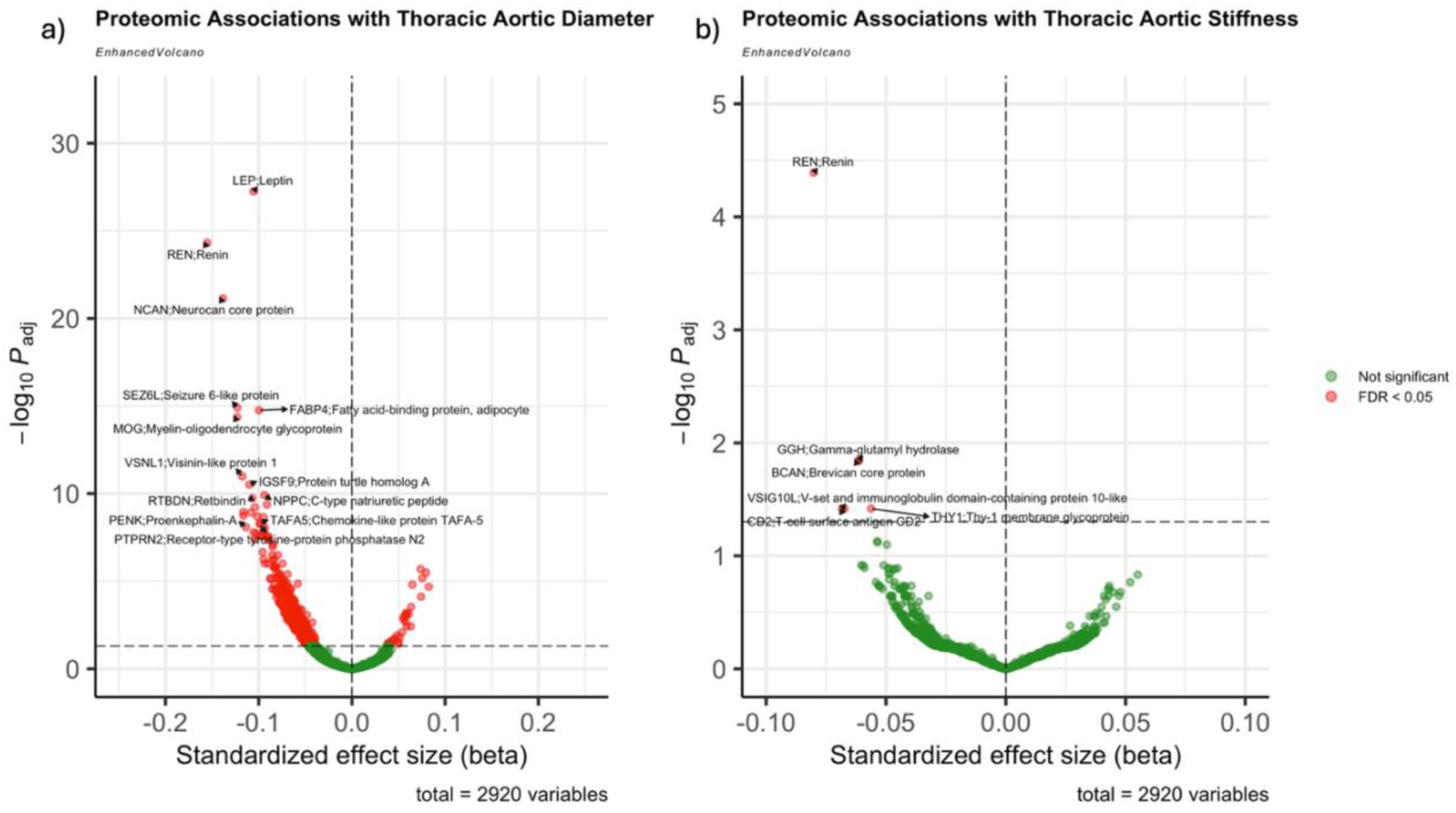
Volcano plot demonstrating proteomic associations with (a) thoracic aortic diameter and (b) thoracic aortic stiffness. The x-axis represents the standardized effect size of the protein, and the y-axis represents the FDR-adjusted p-value.

Sequential adjustment for BMI and SBP altered the number and magnitude of significant protein associations. Addition of BMI resulted in 398 FDR-significant proteins (32 positive; 366 negative relationship), compared with 314 following adjustment for SBP (102 positive; 212 negative relationship). When both BMI and SBP were included, 434 proteins were significant (31 positive; 403 negative relationship), while 417 remained significant (29 positive; 388 negative relationship) following further adjustments for smoking status and ethnicity.

**Figure 3:**
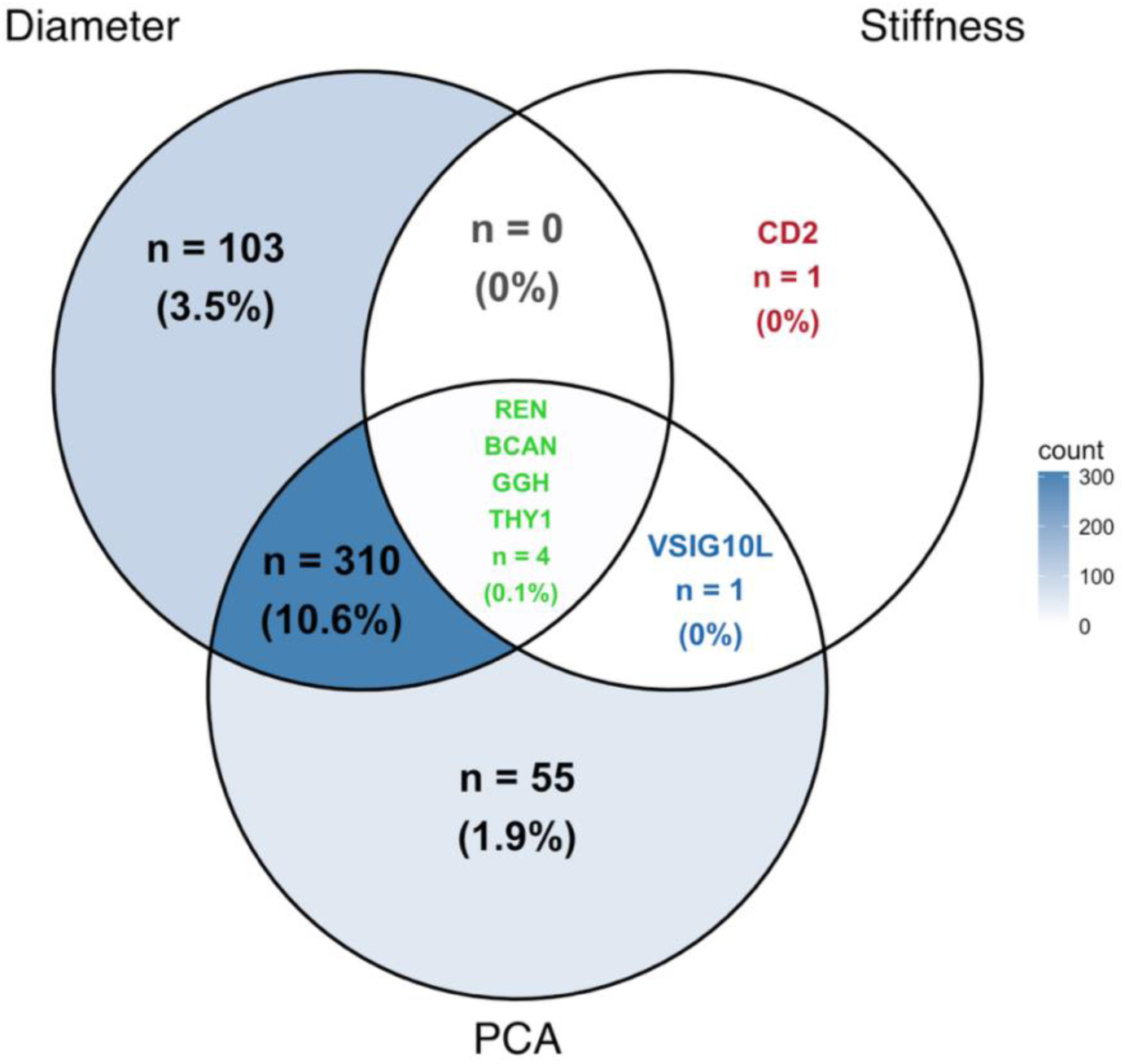
Venn diagram of the three phenotype models (aortic diameter, aortic stiffness and PC1) demonstrating overlaps of significantly associated proteins (FDR < 0.05). The number of proteins and the proportion are represented in each segment. For regions with fewer than 10 significant proteins, individual protein labels are explicitly listed.

### Proteome-wide associations with aortic stiffness

60 proteins demonstrated significant associations (6 positive; 54 negative relationship) with thoracic aortic stiffness in models adjusted for age and sex (Fig. 2b; Supplementary Table 2). The strongest associations included renin (*REN*), gamma-glutamyl hydrolase (*GGH*), fatty acid-binding protein, adipocyte (*FABP4*), galectin-9 (*LGALS9)*, and Thy-1 membrane glycoprotein (*THY1*). Additional adjustment for BMI substantially attenuated proteomic signals, yielding just 6 significantly associated proteins, all of which were inversely associated with increasing stiffness.

The primary fully adjusted model, which additionally included smoking status and ethnicity but excluded SBP, found the same six proteins to remain significantly associated with aortic stiffness. The six proteins following the primary adjustments included: *REN*, brevican core protein (*BCAN*), *GGH*, V-set and immunoglobulin domain-containing protein 10-like (*VSIG10L*), *THY1*, and T-cell surface antigen (*CD2*).

In contrast, sensitivity analyses involving additional adjustment for SBP substantially altered the stiffness-associated proteomic profile, with 296 proteins reaching FDR significance in the age-, sex- and SBP-adjusted model (26 positive; 270 negative relationship). Further adjustment for BMI decreased this number to eight, and additional adjustment for smoking status and ethnicity subsequently identified seven significantly associated proteins; however, these proteins differed substantially from those identified from the primary model, with an overlap between only three of the seven proteins.

### Secondary PCA-derived aortic phenotype proteomic associations

Principal component analysis integrating aortic diameter and stiffness data identified two components; PC1 explained 60.57% and PC2 explained 39.43% of the total phenotypic variance. However, most of the proteins identified from the fully adjusted PC1 phenotype model overlapped with the fully adjusted diameter model, with 310 proteins shared between diameter and PC1. This suggests that the PC1 proteomic profile was driven predominantly by the diameter profile rather than an independent proteomic phenotype.

In the baseline model adjusted for age and sex, 150 of 2920 proteins (18 positive; 132 negative relationship) were significantly associated with PC1 after FDR correction (Supplementary Fig. 1; Supplementary Table 3). The strongest associations included *NCAN*, *SEZ6L*, *REN*, *BCAN*, and *MOG*.

Subsequent covariate adjustment models substantially changed the number of significant PC1 associations. Model 2, further adjusting for BMI, identified 250 FDR-significant proteins (21 positive; 229 negative relationship), while adjustment for SBP identified 191 (11 positive; 180 negative relationship). When both BMI and SBP were adjusted alongside age and sex, 375 proteins were significant (21 positive; 354 negative relationship). In the fully adjusted model further adjusting for smoking status and ethnicity, 370 proteins remained significantly associated, with 20 positively associated and 350 negatively associated (Supplementary Table 3).

In contrast to PC1, PC2 demonstrated opposite loadings for diameter and stiffness (0.707 and −0.707 respectively), representing divergence between structural dilatation and aortic stiffening, whereby dilatation occurs without a similarly proportional increase in stiffness. The proteins associated with the PC2 phenotype can be seen in Supplementary Table 4.

### Overlap between diameter, stiffness and PC1 associations

Overlap analysis suggested substantial number of shared proteins between diameter and the secondary PCA-derived aortic phenotype, with 310 proteins significant in both models. Four proteins were significantly associated across all three phenotypic models, while only one was specific to stiffness alone. No proteins were shared exclusively between the diameter and stiffness models without also having some overlap with PC1. Shared proteins across the three models included *REN*, *BCAN*, *GGH*, and *THY1*.

### Enrichment analysis of significantly associated proteins

Gene set enrichment analysis across the two phenotype models identified patterns of enrichment across the MSigDB hallmark, C2, and Gene Ontology Biological Processes curated pathway collections. Hallmark analysis identified negative enrichment of coagulation and complement pathways with increasing aortic diameter (Fig. 4a). Coagulation had an NES of −1.64, and complement had an NES of −1.46, with both pathways meeting the FDR-adjusted significance threshold. In contrast, neither pathway was significantly enriched for aortic stiffness. Coagulation showed a nonsignificant positive NES (NES = 0.75), whereas complement showed a directionally concordant but nevertheless nonsignificant negative NES (NES = −0.82) for stiffness.

**Figure 4:**
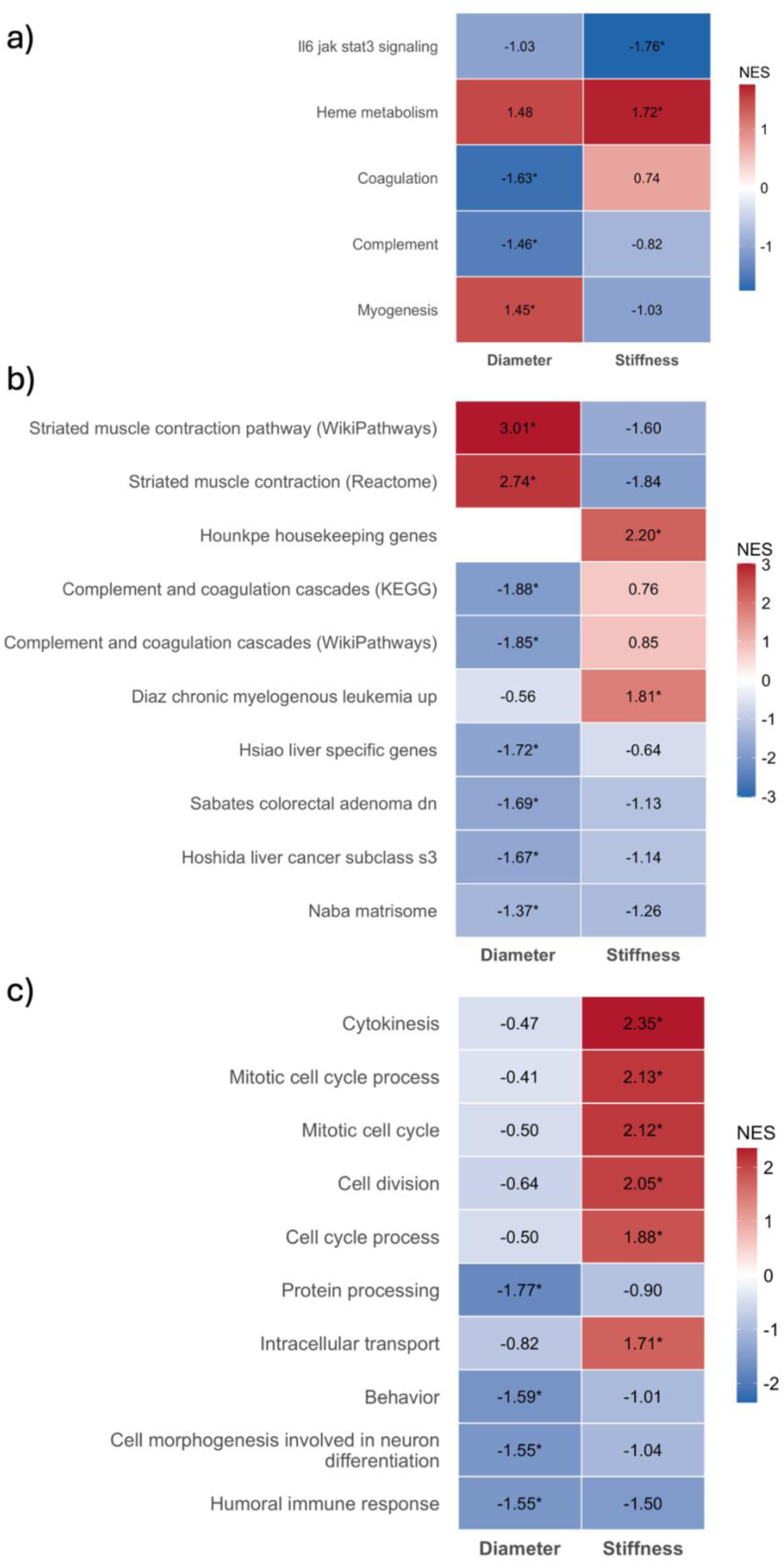
Functional enrichment of proteomic associations across thoracic aortic phenotypes. Gene set enrichment analysis was performed on the complete protein list, ranked according to the signed regression test statistic. Enrichment was assessed using MSigDB (a) Hallmark, (b) Curated Gene Sets (C2), and (c) Gene Ontology Biological Processes (C5). The heat maps show normalized enrichment scores (NES) for pathways reaching an FDR-adjusted p-value < 0.05 in at least one of the two phenotypes, and significant pathways are marked by an asterisk sign. Positive and negative NES values represent enrichment among proteins positively and negatively associated with the corresponding phenotype.

Within the C2 curated pathway collection, 45 pathways were significantly enriched for aortic diameter and 25 for aortic stiffness (Fig. 4b). Diameter-associated pathways included KEGG complement and coagulation cascades (NES = −1.88; FDR = 1.00 × 10^−4^), WP striated muscle contraction pathway (NES = 3.04; FDR = 1.00 × 10^−4^), and NABA matrisome pathway representing extracellular matrix reorganisation (NES = −1.37; FDR = 1.98 × 10^−4^). In contrast, proteins associated with aortic stiffness showed a distinct enrichment pattern characterized by cytokine signalling, glycosaminoglycan synthesis, and EGFR signalling.

Gene Ontology Biological Process analysis identified negatively enriched aortic diameter-associated pathways relating to protein processing, humoral immunity, and cell-junction organization, with positive enrichment of myofibril and sarcomere assembly pathways (Fig. 4c). As with previous gene sets, stiffness showed a more distinct profile, characterized by positively enriched cell-cycle, mitotic and intracellular transport processes, while negatively being enriched for adaptive immune response pathways. Additionally, functional enrichment of the proteins associated with PC1 mostly captured the same pathways as aortic diameter (Supplementary Fig. 2).

Over-representation analysis (ORA) was also performed for larger protein subsets for certain phenotype groups. Diameter-only proteins (n = 103) and PC1-only proteins (n = 55) were not significantly associated with any biological pathways. In contrast, multiple pathways were enriched between diameter and PC1, including humoral immune response and complement activation (specifically the classical pathway). The pathways and the interactions among proteins are visualized in Supplementary Fig. 3. ORA was not conducted for stiffness-specific proteins or proteins shared across all three phenotype models as they contained too few proteins to support inference.

### Candidate protein analysis: *ELN*

Given the well-known role of elastin in aortic wall structure in regulating the diameter and stiffness and its absence in the phenotype models, *ELN* was examined separately (Fig. 5). Circulating *ELN* showed a strong association with age. Although *ELN* was significantly associated with aortic diameter and stiffness in both unadjusted and age-adjusted models (notably for diameter, however, the direction of association reversed from positive to negative after age adjustment), following adjustment of the full covariates, these associations were attenuated (Supplementary Table 5). This suggests that *ELN*-related changes may be largely impacted by age and other cardiovascular risk factors.

**Figure 5:**
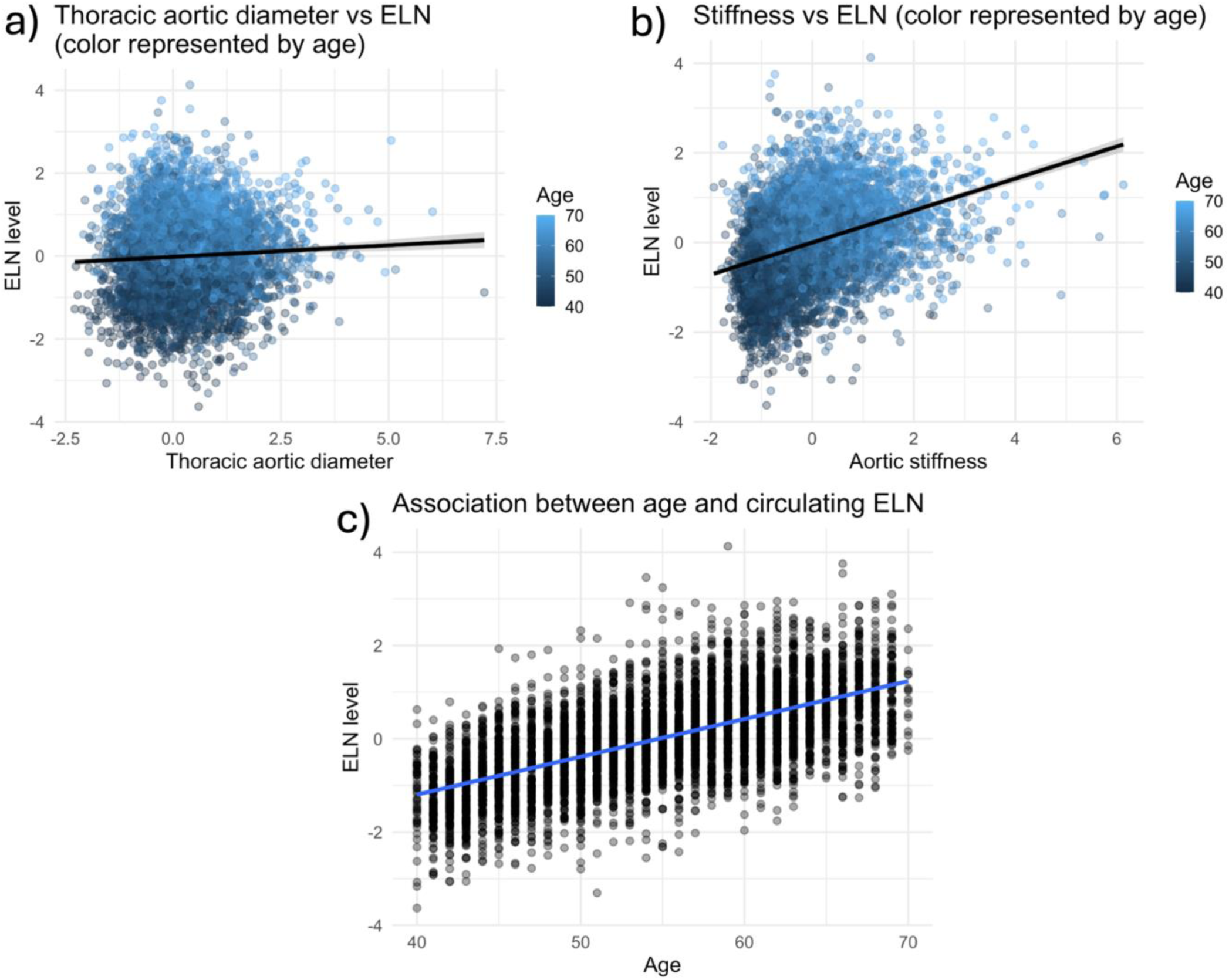
Results of ELN sensitivity analysis. Scatter plots displaying the relationship between circulating ELN levels and (**a**) thoracic aortic diameter and (**b**) thoracic aortic stiffness, with points coloured by patient age. (**c**) Linear regression analysis illustrating the correlation between age and circulating ELN levels.

### Additional secondary analyses

Sex-interaction testing was also conducted, identifying several proteins that demonstrated strong interaction signals with different phenotype models. The sex interactions with diameter and stiffness can be seen in Supplementary Tables 6 and 7, respectively. Among the associated proteins with aortic diameter, tetraspanin-1 (*TSPAN1*), follitropin subunit beta (*FSHB*), glycodelin (*PAEP*), chordin-like protein 2 (*CHRDL2*), and leptin (*LEP*) were the most significantly influenced by sex interactions. *FSHB*, *PAEP*, *TSPAN1*, glycoprotein hormones alpha chain (*CGA*), and receptor-type tyrosine-protein phosphatase zeta (*PTPRZ1*) were among those associated with aortic stiffness that were significantly influenced by sex interactions.

## Discussion

In this cross-sectional proteome-wide association study, aortic diameter and stiffness showed markedly different circulating proteomic profiles. In the baseline age- and sex-adjusted analyses, 358 proteins were associated with diameter and 60 with aortic stiffness following FDR correction. Subsequent additions of covariate adjustments indicated that these associations were sensitive to cardiometabolic covariates, particularly BMI. Across the fully adjusted multivariable linear regression models, aortic diameter was associated with a broad proteomic signature (417 proteins), whereas stiffness was associated with a more restricted proteomic signature (6 proteins). Functional enrichment analyses further revealed differences between the two imaging phenotypes: thoracic aortic diameter was characterized predominantly by complement/coagulation pathways, while stiffness showed a more distinct profile involving cytokinesis, cytoskeletal organization and intracellular transport. Secondary PCA-derived analysis showed that PC1, representing the same-direction component between aortic diameter and stiffness, largely overlapped with the diameter-associated proteomic profile, with 310 out of the 370 proteins being shared in the fully adjusted models.

These findings extend prior genetic, imaging and proteomic work by linking large-scale plasma proteomics to imaging-derived thoracic aortic phenotypes, and by directly comparing the molecular profiles of structural dilation and functional stiffening. Prior work on the UK Biobank dataset has demonstrated that automated cardiac MRI-derived aortic diameter can be used for genetic analysis and predict future TAA risk (13). Our study builds on this by examining whether circulating plasma proteins are associated with continuous thoracic aortic imaging phenotypes, rather than focusing solely on the genetic determinants. Other similar proteomics work conducted in TAA patient populations have been limited to smaller sample sizes and restricted to clinically diagnosed aneurysm events (16,34–37). The current approach may capture earlier biological variation across different aspects of aortic remodelling before patients meet clinical thresholds for a diagnosis of TAA. Current clinical surveillance and intervention decisions remain largely diameter-based, with guideline recommendations using aortic size to classify aneurysmal dilation and determine imaging follow-up (4,5).

From a clinical perspective, diameter and stiffness may capture different aspects or stages of thoracic aortic remodelling. Aortic diameter reflects the structural enlargement of the vasculature and remains the established metric for surveillance and intervention thresholds (4,5). In contrast, stiffness reflects the mechanical properties of the aortic wall and may be influenced by vascular ageing, extracellular matrix composition, and vascular smooth muscle cell function (8). Previous work has suggested that aortic stiffness may act as a potential marker for adverse cardiovascular outcomes and dissection among patients with a dilated ascending aorta (9,10). Therefore, it is possible that aortic diameter and stiffness do not occur synchronously but may instead occur distinctly or temporally separated in the disease progression. The differing proteomic patterns observed in this study are consistent with these findings in the literature, although the cross-sectional design prevents verification of the temporal sequence of events.

Sequential covariate adjustment also found that the observed proteomic associations were substantially influenced by cardiometabolic factors. For aortic diameter, adjustment for BMI substantially attenuated the magnitude of several leading protein associations, whereas additional adjustment for SBP generally produced smaller changes once BMI was accounted for. Further inclusion of both smoking status and ethnicity resulted in comparatively modest additional changes, suggesting that much of the covariate-related variation was accounted for by BMI and SBP. Similarly, substantial attenuation was associated with additional adjustment of BMI in the stiffness phenotype model, with only six of 60 proteins remaining significant. However, incorporating SBP in the sensitivity analyses substantially increased the number of significant protein-phenotype associations. Given the direct inclusion of SBP in the derivation of the stiffness phenotype, the primary stiffness model excluded SBP from adjustment. It is also important to note that the observed impact of the covariates may not necessarily be evidence of confounding alone; BMI and blood pressure may also act as mediators linking circulating proteins to aortic remodelling, which this cross-sectional design cannot distinguish.

A key finding was the biological difference underlying aortic diameter and stiffness changes. Aortic diameter was associated with hundreds of proteins in the fully adjusted model, with most showing inverse associations (388/417 proteins), whereas stiffness had a much more restricted molecular signature (six proteins) in the primary fully adjusted model. This suggests that aortic diameter and stiffness should not be treated as interchangeable measures of the same disease process. The broader diameter-associated signature seems to reflect more extensive changes affecting various types of proteins, while stiffness may capture a more restricted phenotype reflecting weaker effect sizes, greater variability between patients, differences in biological processes between diameter and stiffness, or any combination of these factors. The secondary PCA analysis found mostly similar diameter-associated signals; however, 55 proteins were uniquely associated with PC1, suggesting that combining the two aortic phenotypes may capture additional proteomic signal not detected by either alone.

Complement and coagulation pathways represented one of the most consistent signals associated with aortic diameter. Negative enrichment was observed across Hallmark, C2 curated pathways and GO Biological Processes collections, indicating that these proteins tended to show inverse associations with increasing diameter. Although complement and coagulation pathways have previously been implicated as activated processes in aneurysmal biology, contributing to aortic medial degeneration through regulation of matrix metalloproteinases, the functional enrichment analysis from this present study identified negative enrichment with increasing aortic diameter (38,39). This may reflect potential differences between systemic levels of proteins and local aortic wall activity. Complement factors, such as C3b, have been shown to deposit within the aortic wall (40), alongside increased expression of complement receptors (38), potentially contributing to local consumption of the proteins. A proteomic analysis of TAA at both tissue and plasma levels has also identified complement and coagulation cascades across both scales, supporting their involvement. Additionally, this may reflect important methodological distinctions, with previous studies primarily having examined complement activation within aneurysmal tissue models or patients with established thoracic aortic disease (38–40), whereas the present analysis evaluated circulating plasma proteins across a wide continuum of imaging phenotypes.

Diameter was additionally enriched for extracellular-matrix and contractile pathways, including matrisome, ECM regulators, and sarcomere organization. These findings are biologically consistent with the pathophysiology of TAA, characterized by medial degeneration, extracellular-matrix disruption, and vascular smooth muscle dysfunction (41–44). Other proteomics analyses have also previously shown similar findings (34).

In contrast, although only 6 proteins were associated with stiffness after FDR correction and covariate adjustment, GSEA of the ranked proteome showed a more distinct enrichment profile. GO Biological Process analysis highlighted cell-cycle, mitotic, cytokinesis, cytoskeletal organisation and intracellular transport pathways, while the C2 collection enrichment suggested cytokine signalling, glycosaminoglycan synthesis and EGFR signalling. This is consistent with functional enrichment analyses of proteins identified in patients with aortic aneurysm (45,46). However, considering only six proteins reached FDR significance for stiffness, the findings from stiffness may reflect weaker protein-level effects, and require validated in other independent cohorts.

The limited overlap across the two phenotype models is biologically informative and may help prioritize candidate circulating biomarkers for future biomarker investigation. *REN*, *BCAN*, *GGH* and *THY1* were the only proteins significantly associated with diameter, stiffness and the secondary PC1 phenotype in the fully adjusted models. Notably, *REN* is known to be important in the renin-angiotensin-aldosterone system (RAAS) in blood pressure regulation and vascular remodelling (47). *BCAN* is a type of proteoglycan, belonging to the lectican family, members of which have been implicated in proteoglycan accumulation and medial degeneration in TAA (48). Additionally, *THY1* may reflect immune or cell adhesion pathways when responding to paracrine signals to promote proliferation and vasculogenesis (49,50). Overall, the small number of overlapping proteins suggests that thoracic aortic remodelling involves partly overlapping but mostly phenotype-specific molecular processes.

The observed protein-phenotype associations may have several explanations, including a causal effect of circulating proteins on aortic remodelling, residual or shared confounding, or reverse causation whereby changes in aortic function influence circulating protein levels. Given the cross-sectional nature of this study, these possibilities cannot be distinguished.

From a translational perspective, an important future direction will be to determine whether the circulating proteomic signatures identified in this study can provide incremental predictive value beyond the established imaging-derived aortic diameter and risk factors (13,51). Multimodal risk models combining established variables with selected circulating proteins should be evaluated longitudinally for their ability to predict incident TAA. Such models may also help determine at what stage proteomic changes associated with aortic stiffness occurs relative to those accompanying structural dilation, thereby clarifying whether these phenotypes capture different stages of disease progression. Conversely, establishing individual proteins as causal mediators or therapeutic targets would require complementary genetic approaches, such as Mendelian randomisation, experimental validation, and ultimately interventional clinical studies.

Exploratory sex-interaction analyses additionally suggested that some protein-phenotype associations may differ between male and female participants. Notably, among these proteins, *LEP* demonstrated a stronger inverse relationship with aortic diameter and stiffness in females than males, and *BCAN* demonstrated a greater magnitude of effect on aortic stiffness in men. However, these analyses were exploratory, and future studies should investigate how sex modifies proteomic signatures of aortic remodelling, particularly given the known impact of sex on thoracic aortic aneurysm progression and outcomes (52–54).

One limitation of this present study is its cross-sectional design, which limits inference about temporality or causality. Further Mendelian randomisation analyses based on an external GWAS dataset or other causal-inference approaches may help assess whether these proteins have a causal role. Related to this, plasma proteomic samples and cardiac imaging were collected at different UK Biobank visits. Although one analysis integrating UK Biobank proteomics with multi-organ imaging found that this temporal gap did not have a significant impact on the associations (22), the possibility that circulating protein levels changed between the visits could not nevertheless be excluded. The sequential covariate analyses cannot determine whether changes in protein-phenotype associations following adjustment for BMI and SBP reflect minimizing confounding effects or adjustment for factors lying on the causal pathway. Second, the level of circulating proteins may not necessarily reflect aortic tissue-specific changes and may reflect other systemic changes. Third, the UK Biobank contains mostly volunteers who are predominantly white (96.07%) and healthier than the general population, limiting generalizability (55). Fourth, Olink proteomics measures a predefined panel of proteins rather than the complete list of circulating proteins. Fifth, this study did not adjust for kidney function (eGFR). Renin, in particular, is influenced by kidney function and other determinants of the renin-angiotensin-aldosterone system (56); it is possible that the observed association is partially reflected by these residual confounders rather than aortic biology.

## Conclusion

Circulating proteomic associations with thoracic aortic remodelling differed considerably between structural dilation and functional stiffening and were influenced by adjustment for cardiometabolic factors. Aortic diameter was characterized by complement-coagulation, extracellular matrix, and contractile signatures, whereas stiffness showed a smaller but more distinct pattern involving cytokine and proliferative signalling, cell cycle regulation, and intracellular transport. Secondary PCA analysis suggested that the same-direction component between diameter and stiffness largely reflected the diameter-associated proteomic profile. These findings suggest partly overlapping but differing proteomic association patterns underlying thoracic aortic remodelling across structural dilation and functional stiffening. Proteins associated with both aortic dilation and stiffness may provide a starting point for biomarker discovery, although longitudinal and external validation are further required.

## Sources of Funding

NP is supported by the UCL BHF Research Accelerator AA/18/6/34223, the National Institute for Health and Care Research University College London Hospitals Biomedical Research Centre (NIHR UCLH BRC), and the Aortic Dissection Charitable Trust. JT and ADH are also supported by the NIHR UCLH BRC. UCL receives additional support from a BHF Centre of Research Excellence award. AJH is supported by a BHF Programme Grant (RG/F/23/110123).

## Disclosures

AJH is a scientific advisory board member/consultant for or has received research funding from Ascendis Pharma A/S, BioMarin Pharmaceutical, PharmaIN Corp., Bionevix Ltd. and Novo Nordisk. The other authors declare no conflicts of interest.

## Acknowledgements

This research was conducted using the UK Biobank Resource under Application Number 12113. We thank the UK Biobank participants and the UK Biobank team.

## Author Contributions

AC performed the data collection, analysis and drafted the manuscript. AJH, JT, and ADH critically revised the manuscript. NP conceived the study, supervised the work, and drafted the manuscript. All authors read and approved the final version.

## Data Availability

UK Biobank data are available to approved researchers through application. Full protein-level results are provided in Supplementary Tables 1–7. Further data will be made available upon reasonable request.

## Notes

### Author Declarations

The North West Multi-centre Research Ethics Committee gave ethical approval for the UK Biobank study (21/NW/0157), under which this research was conducted. All participants provided informed consent.

## References

1. Johansson G, Markström U, Swedenborg J. Ruptured thoracic aortic aneurysms: A study of incidence and mortality rates. J Vasc Surg. 1995 Jun;21(6):985–8. doi:10.1016/S0741-5214(95)70227-X

2. Gouveia E, Melo R, Silva Duarte G, Lopes A, Alves M, Caldeira D, Fernandes E, Fernandes R, et al. Incidence and Prevalence of Thoracic Aortic Aneurysms: A Systematic Review and Meta-analysis of Population-Based Studies. Semin Thorac Cardiovasc Surg. 2022;34(1):1–16. doi:10.1053/j.semtcvs.2021.02.029

3. Ying AJ, Affan ET. Abdominal Aortic Aneurysm Screening: A Systematic Review and Meta-analysis of Efficacy and Cost. Ann Vasc Surg. 2019 Jan;54:298–303.e3. doi:10.1016/j.avsg.2018.05.044

4. Isselbacher EM, Preventza O, Hamilton Black J, Augoustides JG, Beck AW, Bolen MA, et al. 2022 ACC/AHA Guideline for the Diagnosis and Management of Aortic Disease: A Report of the American Heart Association/American College of Cardiology Joint Committee on Clinical Practice Guidelines. Circulation. 2022 Dec 13;146(24). doi:10.1161/CIR.0000000000001106

5. Mazzolai L, Teixido-Tura G, Lanzi S, Boc V, Bossone E, Brodmann M, et al. 2024 ESC Guidelines for the management of peripheral arterial and aortic diseases. Eur Heart J. 2024 Sep 29;45(36):3538–700. doi:10.1093/eurheartj/ehae179

6. Proietti R, Field M, McKay V, Lip GYH, Kuduvalli M. Screening for the vulnerable aorta: targeting high-risk groups in the population. Br J Cardiol. 2023;30(3):25. doi:10.5837/bjc.2023.025 PubMed PMID: 39144089; PubMed Central PMCID: PMC11321460.

7. Persson SE, Holsti M, Mani K, Wanhainen A. Difference in risk factor profile for abdominal aortic aneurysm and thoracic aortic aneurysm. J Vasc Surg. 2025 Feb;81(2):335–341.e6. doi:10.1016/j.jvs.2024.10.012

8. Herzog MJ, Müller P, Lechner K, Stiebler M, Arndt P, Kunz M, et al. Arterial stiffness and vascular aging: mechanisms, prevention, and therapy. Signal Transduct Target Ther. 2025 Sep 1;10(1):282. doi:10.1038/s41392-025-02346-0

9. Rosnel C, Sivera R, Cervi E, Danton M, Schievano S, Capelli C, et al. Are aortic biomechanical properties early markers of dilatation in patients with Marfan syndrome? A systematic review and meta-analysis. Biomech Model Mechanobiol. 2024 Dec;23(6):2043–61. doi:10.1007/s10237-024-01881-z

10. Fortunato RN, Huckaby LV, Emerel LV, Schlosser V, Yang F, Phillippi JA, et al. The predictive capability of aortic stiffness index for aortic dissection among dilated ascending aortas. J Thorac Cardiovasc Surg. 2024 Jun;167(6):2015–24. doi:10.1016/j.jtcvs.2022.09.003

11. Singh B, Karpov OA, Mayr M. Clinical proteomics in cardiovascular medicine: Current capabilities, limitations, and future directions. Atherosclerosis. 2026 Feb;413:120637. doi:10.1016/j.atherosclerosis.2026.120637

12. Lam MPY, Ping P, Murphy E. Proteomics Research in Cardiovascular Medicine and Biomarker Discovery. J Am Coll Cardiol. 2016 Dec;68(25):2819–30. doi:10.1016/j.jacc.2016.10.031

13. Pirruccello JP, Chaffin MD, Chou EL, Fleming SJ, Lin H, Nekoui M, et al. Deep learning enables genetic analysis of the human thoracic aorta. Nat Genet. 2022 Jan;54(1):40–51. doi:10.1038/s41588-021-00962-4

14. Tcheandjieu C, Xiao K, Tejeda H, Lynch JA, Ruotsalainen S, Bellomo T, et al. High heritability of ascending aortic diameter and trans-ancestry prediction of thoracic aortic disease. Nat Genet. 2022 Jun;54(6):772–82. doi:10.1038/s41588-022-01070-7

15. Lemoli M, Renc P, Agabiti Rosei C, De Ciuceis C, Nosalski R, Sitek A, et al. UK Biobank plasma proteomic data identify predictors of arterial stiffness and high blood pressure. Eur Heart J. 2025 Nov 5;46(Supplement_1):ehaf784.3315. doi:10.1093/eurheartj/ehaf784.3315

16. Daskalopoulou A, Giotaki SG, Toli K, Minia A, Pliaka V, Alexopoulos LG, et al. Targeted Proteomic Analysis of Patients with Ascending Thoracic Aortic Aneurysm. Biomedicines. 2023 Apr 25;11(5):1273. doi:10.3390/biomedicines11051273

17. Sudlow C, Gallacher J, Allen N, Beral V, Burton P, Danesh J, et al. UK biobank: an open access resource for identifying the causes of a wide range of complex diseases of middle and old age. PLoS Med. 2015 Mar;12(3):e1001779. doi:10.1371/journal.pmed.1001779 PubMed PMID: 25826379; PubMed Central PMCID: PMC4380465.

18. Chong SH, Saccoh AF, Van Vugt M, Schmidt AF, Paliwal N. Integrative Observational and Genetic Evidence From the UK Biobank Supports Carotid Intima-Media Thickness as a Window Into Cardiac Remodeling. J Am Heart Assoc. 2026 Jul 7;15(13):e049139. doi:10.1161/JAHA.125.049139

19. Bai W, Sinclair M, Tarroni G, Oktay O, Rajchl M, Vaillant G, et al. Automated cardiovascular magnetic resonance image analysis with fully convolutional networks. J Cardiovasc Magn Reson. 2018 Feb;20(1):65. doi:10.1186/s12968-018-0471-x

20. Petersen SE, Matthews PM, Francis JM, Robson MD, Zemrak F, Boubertakh R, et al. UK Biobank’s cardiovascular magnetic resonance protocol. J Cardiovasc Magn Reson. 2016 Jan;18(1):8. doi:10.1186/s12968-016-0227-4

21. Paliwal N, Henry A, Finan C, Davies RH, Lumbers RT, Tsui J, et al. Development and evaluation of Z-score based aortic diameter thresholds for early detection of thoracic aortic dissection and aneurysm: Analysis in the UK Biobank [Internet]. Cardiovascular Medicine; 2025 [cited 2026 Jul 16]. Available from: http://medrxiv.org/lookup/doi/10.1101/2025.10.03.25337259 doi:10.1101/2025.10.03.25337259

22. Fan Z, Chirinos J, Yang X, Shu J, Li Y, O’Brien JM, et al. The landscape of plasma proteomic links to human organ imaging. Nat Commun. 2026 Jul 11;17(1):8565. doi:10.1038/s41467-026-74715-4

23. Chen ML, Kho PF, Guarischi-Sousa R, Zhou J, Panyard DJ, Azizi Z, et al. Plasma proteomics and carotid intima-media thickness in the UK biobank cohort. Front Cardiovasc Med. 2024 Oct 2;11:1478600. doi:10.3389/fcvm.2024.1478600

24. Cecelja M, Ruijsink B, Puyol-Antón E, Li Y, Godwin H, King AP, et al. Aortic Distensibility Measured by Automated Analysis of Magnetic Resonance Imaging Predicts Adverse Cardiovascular Events in UK Biobank. J Am Heart Assoc. 2022 Dec 6;11(23):e026361. doi:10.1161/JAHA.122.026361

25. Paliwal N, Henry A, Finan C, Davies R, Hughes A, Williams B, et al. Abstract 4141107: Genome-wide association study of aortic stiffness derived from deep learning in CMR images of 45,789 individuals identifies loci linked with cell-matrix structure. Circulation. 2024 Nov 12;150(Suppl_1). doi:10.1161/circ.150.suppl_1.4141107

26. Barrett T, Dowle M, Srinivasan A, Gorecki J, Chirico M, Hocking T, et al. data.table: Extension of ‘data.framè [Internet]. 2006 [cited 2026 Jul 16]. p. 1.18.4. Available from: https://CRAN.R-project.org/package=data.table doi:10.32614/CRAN.package.data.table

27. Carrasco-Zanini J, Wheeler E, Uluvar B, Kerrison N, Koprulu M, Wareham NJ, et al. Mapping biological influences on the human plasma proteome beyond the genome. Nat Metab. 2024 Sep 26;6(10):2010–23. doi:10.1038/s42255-024-01133-5

28. Koprulu M, Wheeler E, Kerrison ND, Denaxas S, Carrasco-Zanini J, Orkin CM, et al. Sex differences in the genetic regulation of the human plasma proteome. Nat Commun. 2025 May 13;16(1):4001. doi:10.1038/s41467-025-59034-4

29. Benjamini Y, Hochberg Y. Controlling the False Discovery Rate: A Practical and Powerful Approach to Multiple Testing. J R Stat Soc Ser B Stat Methodol. 1995 Jan 1;57(1):289–300. doi:10.1111/j.2517-6161.1995.tb02031.x

30. Wu T, Hu E, Xu S, Chen M, Guo P, Dai Z, et al. clusterProfiler 4.0: A universal enrichment tool for interpreting omics data. The Innovation. 2021 Aug;2(3):100141. doi:10.1016/j.xinn.2021.100141

31. Carlson M. org.Hs.eg.db [Internet]. Bioconductor; 2017 [cited 2026 Jul 16]. Available from: https://bioconductor.org/packages/org.Hs.eg.db doi:10.18129/B9.BIOC.ORG.HS.EG.DB

32. Karimi A, Milewicz DM. Structure of the Elastin-Contractile Units in the Thoracic Aorta and How Genes That Cause Thoracic Aortic Aneurysms and Dissections Disrupt This Structure. Can J Cardiol. 2016 Jan;32(1):26–34. doi:10.1016/j.cjca.2015.11.004 PubMed PMID: 26724508; PubMed Central PMCID: PMC4839280.

33. Shen C, Zhang R, Yu J, Sahakian BJ, Cheng W, Feng J. Plasma proteomic signatures of social isolation and loneliness associated with morbidity and mortality. Nat Hum Behav. 2025 Jan 3;9(3):569–83. doi:10.1038/s41562-024-02078-1

34. Serhatli M, Baysal K, Acilan C, Tuncer E, Bekpinar S, Baykal AT. Proteomic Study of the Microdissected Aortic Media in Human Thoracic Aortic Aneurysms. J Proteome Res. 2014 Nov 7;13(11):5071–80. doi:10.1021/pr5006586

35. Matsumoto KI, Maniwa T, Tanaka T, Satoh K, Okunishi H, Oda T. Proteomic analysis of calcified abdominal and thoracic aortic aneurysms. Int J Mol Med. 2012 Aug;30(2):417–29. doi:10.3892/ijmm.2012.985

36. Tyrrell DJ, Chen J, Li BY, Wood SC, Rosebury-Smith W, Remmer HA, et al. Aging Alters the Aortic Proteome in Health and Thoracic Aortic Aneurysm. Arterioscler Thromb Vasc Biol. 2022 Aug;42(8):1060–76. doi:10.1161/ATVBAHA.122.317643

37. Kazamia R, Keravnou A, Moushi A, Sokratous K, Michailidou K, Yiangou K, et al. Tissue and plasma proteomic profiling indicates AHSG as a potential biomarker for ascending thoracic aortic aneurysms. BMC Cardiovasc Disord. 2023 Mar 16;23(1):138. doi:10.1186/s12872-023-03154-6

38. Piao C, Zhang WM, Deng J, Zhou M, Liu TT, Zheng S, et al. Activation of the alternative complement pathway modulates inflammation in thoracic aortic aneurysm/dissection. Am J Physiol-Cell Physiol. 2024 Feb 1;326(2):C647–58. doi:10.1152/ajpcell.00210.2023

39. Fiane AE, Videm V, Lingaas PS, Heggelund L, Nielsen EW, Geiran OR, et al. Mechanism of Complement Activation and Its Role in the Inflammatory Response After Thoracoabdominal Aortic Aneurysm Repair. Circulation. 2003 Aug 19;108(7):849–56. doi:10.1161/01.CIR.0000084550.16565.01

40. Zhang F, Yao K, Liu Y, Zhou M, Zhang Y, Hong S, et al. Complement C3a/C3aR inhibition alleviates the formation of aortic aneurysm in Marfan syndrome mice. BMC Cardiovasc Disord. 2024 Aug 10;24(1):417. doi:10.1186/s12872-024-04077-6 PubMed PMID: 39127656; PubMed Central PMCID: PMC11316375.

41. Jana S, Hu M, Shen M, Kassiri Z. Extracellular matrix, regional heterogeneity of the aorta, and aortic aneurysm. Exp Mol Med. 2019 Dec 19;51(12):1–15. doi:10.1038/s12276-019-0286-3 PubMed PMID: 31857579; PubMed Central PMCID: PMC6923362.

42. Mimler T, Nebert C, Eichmair E, Winter B, Aschacher T, Stelzmueller ME, et al. Extracellular matrix in ascending aortic aneurysms and dissections - What we learn from decellularization and scanning electron microscopy. PloS One. 2019;14(3):e0213794. doi:10.1371/journal.pone.0213794 PubMed PMID: 30883576; PubMed Central PMCID: PMC6422325.

43. Cho MJ, Lee MR, Park JG. Aortic aneurysms: current pathogenesis and therapeutic targets. Exp Mol Med. 2023 Dec 1;55(12):2519–30. doi:10.1038/s12276-023-01130-w

44. Rombouts KB, Van Merrienboer TAR, Ket JCF, Bogunovic N, Van Der Velden J, Yeung KK. The role of vascular smooth muscle cells in the development of aortic aneurysms and dissections. Eur J Clin Invest. 2022 Apr;52(4):e13697. doi:10.1111/eci.13697

45. Li M, He X, Gong W, Xiao S, Fu K, Qin Q, et al. Plasma proteomics profiles predict the risk of future aortic aneurysm and aortic dissection. Int J Surg. 2025 Oct 1;111(10):6894–904. doi:10.1097/JS9.0000000000002845 PubMed PMID: 40576182; PubMed Central PMCID: PMC12527807.

46. Bashore AC, Radkevich E, Roudko V, Chen Z, Nie K, Rizakos T, et al. Integrative Omics Identifies Candidate Plasma Biomarkers and Cellular Targets Associated with Thoracic Aortic Aneurysm [Internet]. Cell Biology; 2025 [cited 2026 Jul 16]. Available from: http://biorxiv.org/lookup/doi/10.1101/2025.07.25.666909 doi:10.1101/2025.07.25.666909

47. Pacurari M, Kafoury R, Tchounwou PB, Ndebele K. The Renin-Angiotensin-aldosterone system in vascular inflammation and remodeling. Int J Inflamm. 2014;2014:689360. doi:10.1155/2014/689360 PubMed PMID: 24804145; PubMed Central PMCID: PMC3997861.

48. Cikach FS, Koch CD, Mead TJ, Galatioto J, Willard BB, Emerton KB, et al. Massive aggrecan and versican accumulation in thoracic aortic aneurysm and dissection. JCI Insight. 2018 Mar 8;3(5):e97167. doi:10.1172/jci.insight.97167

49. Yang J, Zhan XZ, Malola J, Li ZY, Pawar JS, Zhang HT, et al. The multiple roles of Thy-1 in cell differentiation and regeneration. Differentiation. 2020 May;113:38–48. doi:10.1016/j.diff.2020.03.003

50. He D, Mao A, Zheng CB, Kan H, Zhang K, Zhang Z, et al. Aortic heterogeneity across segments and under high fat/salt/glucose conditions at the single-cell level. Natl Sci Rev. 2020 May 1;7(5):881–96. doi:10.1093/nsr/nwaa038

51. Henry M, Campello Jorge CA, Van Bakel PAJ, Knauer HA, MacEachern M, Van Herwaarden JA, et al. Thoracic Aortic Aneurysm Growth Rates and Predicting Factors: A Systematic Review and Meta-Analysis. J Am Heart Assoc. 2025 Apr;14(7):e038821. doi:10.1161/JAHA.124.038821

52. Han X, Ding J, Feng S, Li Y, Gao Y, Sun R. Sex-specific prediction models for aortic aneurysm integrating traditional clinical risk factors and proteomic profiles: a large-scale prospective study from the UK Biobank. Biol Sex Differ. 2026 May 11;17(1):128. doi:10.1186/s13293-026-00913-w

53. Jiang H, Xu H, Xu Z. Sex-related differences in outcome of thoracic aortic surgery. J Cardiothorac Surg. 2024 Apr 16;19(1):226. doi:10.1186/s13019-024-02735-6 PubMed PMID: 38627818; PubMed Central PMCID: PMC11020790.

54. Cheung K, Boodhwani M, Chan KL, Beauchesne L, Dick A, Coutinho T. Thoracic Aortic Aneurysm Growth: Role of Sex and Aneurysm Etiology. J Am Heart Assoc. 2017 Feb 3;6(2):e003792. doi:10.1161/JAHA.116.003792 PubMed PMID: 28159818; PubMed Central PMCID: PMC5523737.

55. Fry A, Littlejohns TJ, Sudlow C, Doherty N, Adamska L, Sprosen T, et al. Comparison of Sociodemographic and Health-Related Characteristics of UK Biobank Participants With Those of the General Population. Am J Epidemiol. 2017 Nov 1;186(9):1026–34. doi:10.1093/aje/kwx246

56. Brewster UC, Perazella MA. The renin-angiotensin-aldosterone system and the kidney: effects on kidney disease. Am J Med. 2004 Feb;116(4):263–72. doi:10.1016/j.amjmed.2003.09.034

